# A Novel Laboratory-Developed Test Using Multiplex qPCR to Further Personalize Tacrolimus Dosing

**DOI:** 10.64898/2026.05.30.26354529

**Authors:** Abhishek Chadha, Zhiwei Wang, Max Mamroth, John Hunter, Lin Xu, Sanghamitra Sahoo, Marc Rumpler, Alexandre Vlassov, Anna Chikova

## Abstract

Tacrolimus is an immunosuppressant drug commonly used in transplantation. Although multiple studies have demonstrated that polymorphisms in the *CYP3A5* gene impact the metabolism of tacrolimus, routine pre-transplant testing for these markers is still not broadly implemented. TacroType™ – a new laboratory developed test implemented by One Lambda Laboratories – utilizes a qPCR-based six-plex assay for *CYP3A5* genotyping and detects the three most common genetic variants (*3, *6 and *7) associated with loss of CYP3A5 protein function and reduced tacrolimus metabolism. TacroType was optimized to address known sources of protocol, technical or sample variability to achieve accurate and reproduceable genotyping results.

An analytical performance study was completed following CLSI guidelines. Accuracy was confirmed for each possible *CYP3A5* genotype involving 6 target alleles using 32 well-characterized reference samples. TacroType exhibited accurate performance within a broad range of DNA concentrations and quality. Precision studies indicated consistent genotyping results across 4 operators, 2 instrument types and 5 lots of reagents. Accurate and reproducible assay performance was demonstrated using whole blood from 100 and buccal swabs from 70 donors.

The analytical performance of TacroType was evaluated in 4014 total qPCR reactions, with a report rate of 99.8% and genotyping accuracy of 100% (95% confidence interval of 99.9%).

## 1. Introduction

Tacrolimus is the most commonly utilized induction immunosuppressive medication that is prescribed in kidney[1], pancreas[2], liver[3], intestine[4], heart[5],hematopoietic stem cell[6] and lung[7] transplants. Like cyclosporine[8], tacrolimus functions via the inhibition of calcineurin, a phosphatase that impacts T-cell proliferation[9]. Tacrolimus has a narrow therapeutic window, with either acute rejection[10,11] or acute kidney injury[12] resulting from blood tacrolimus levels that are too low or too high, respectively. Tacrolimus is a substrate for the CYP3A5 enzyme in the liver and gastrointestinal tract[13], and approximately 50% of individual variability in tacrolimus metabolism is attributable to *CYP3A5* genotype[14].

Pharmacokinetic studies have demonstrated that CYP3A5 activity level, commonly defined in scientific literature as expression status, affects the rate of oral tacrolimus clearance, defined as the volume of blood cleared of tacrolimus per unit time following oral administration of the drug[15,16]. The ratio of expressor to non-expressor oral clearance includes estimates of 1.45-fold[17], 1.6-fold[18], 1.68-fold[19], 1.9-fold[16], 2.1-fold[20] and 2.4-fold[21]. For liver transplants, oral tacrolimus clearance depends on the genotype of both the recipient and the donor; clearance was 1.7-fold for donor/recipient expressors compared to donor/recipient non-expressors, while a non-expressor liver grafted onto an expressor liver resulted in a 1.3-fold higher rate of clearance[22]. The relationship between tacrolimus dosing requirements and *CYP3A5* genotype has been demonstrated for multiple transplant types, including lung[23], heart[24], kidney[25] and hematopoietic stem cell transplants[26], for adult as well as pediatric[27] patients.

Several organizations, including the Food and Drug Administration (FDA)[28], Clinical Pharmacogenetics Implementation Consortium (CPIC)[29], Dutch Pharmacogenetics Working Group (DPWG)[30] and Association for Molecular Pathology (AMP)[31] disclose information about the role of CYP3A5 in tacrolimus metabolism. CPIC recommends 1.5 – 2 fold increase of starting tacrolimus dosage for patients with one or two functional copies of *CYP3A5*[29].

Multiple clinical trials have tested whether *CYP3A5*-guided tacrolimus dosing enables patients to reach target blood tacrolimus levels more quickly than standard weight-based method. A prospective, randomized 2-arm clinical trial demonstrated that 54.8% of patients dosed using genotyping data achieved target tacrolimus levels after their first dose, as opposed to 20.8% of patients dosed using a standard method based on weight alone [32]. A similar study found that genotype-guided dosing resulted in 40.3% of genotype-guided patients achieving target tacrolimus levels on day 3 post-transplant, versus 23.8% of weight-guided patients[33]. A one-armed trial utilizing *CYP3A5* to guide tacrolimus dosing found that 58% of patients achieved target tacrolimus levels at day 3 post-transplant, which was expected to be higher than the proportion of patients historically achieving target blood levels of tacrolimus with weight-based methods[34]. Similar results were observed for pediatric patients, for whom *CYP3A5*-guided tacrolimus dosing resulted in faster attainment of target therapeutic tacrolimus concentrations than the standard dosing arm[35]. In contrast, a prospective clinical trial by Shuker et al., 2016, that was performed with kidney transplant recipients found no difference in the proportion of patients with tacrolimus in the target range between genotype-guided and standard protocol groups. However, it is important to note that the genotyping method used in that study did not detect two *CYP3A5* alleles, *6 and *7, which are associated with loss of protein function and are relatively common among individuals of African descent, despite a significant portion of participants in that study being individuals of African ancestry [36]. Observed inconsistency amongst published data may thus result from incomplete genotyping for some study participants.

Improvement of *CYP3A5* genotyping approaches is particularly important for ethnically diverse populations[37]. Incomplete genotyping of individuals of African descent was used in at least three clinical trials assessing the relationship between *CYP3A5* and tacrolimus dosing, and risks dosing patients with toxic supratherapeutic doses of the drug[36,38,39]. African Americans constitute approximately one-third of United States kidney transplant recipients and have lower long-term survival rates than Caucasians or Asians[40]. The FDA label for Prograf® recommends higher dosage for African Americans due to observed racial pharmacokinetic differences[41]. These pharmacokinetic differences are now associated with the relatively higher frequency of fully functioning CYP3A5 in individuals of African ancestry, compared to other ethnic groups[37,42,43]. Nevertheless, multiple studies suggest that genetic analysis of *CYP3A5* can provide better guidance for tacrolimus dose selection than ethnicity alone.

Several studies have linked *CYP3A5* testing with tangible benefits. Consistent with a need for differing initial dosage guidelines, retrospective analysis indicates that *CYP3A5* expressors required significantly more dosage adjustments than *CYP3A5* non-expressors[44]. Accordingly, genotype-guided dosing for tacrolimus has been demonstrated to result in fewer dosage adjustments for both immediate-release[45] as well as extended-release[46] formulations of tacrolimus. *CYP3A5* testing is predicted to mitigate patient costs by reducing the time required to achieve target tacrolimus concentration[47]. Pharmacokinetic modeling studies demonstrate a higher proportion of allogenic-hematopoietic stem cell patients achieving target tacrolimus using genotype-guided dosing versus a purely weight-based protocol[48]. Pharmacogenetic testing of *CYP3A5* has been demonstrated to reduce costs, hospitalization days and the risk of adverse events in a cohort of Austrian patients[49]. As a result of these benefits, transplant centers are actively considering *CYP3A5* testing protocols for tacrolimus dose adjustments[50].

To perform *CYP3A5* genotyping in ethnically diverse populations, 6 separate alleles must be assessed, requiring either several 1-2-plex qPCR reactions or a single 6-plex assay to obtain all required information. TacroType targets both alleles for rs776746, rs10264272 and rs41303343 polymorphic sites (SNPs)[31], also described as loci for the *CYP3A5* *3, *6 and *7 variants. *CYP3A5* *1> *3 in rs776746 corresponds to an A>G (T>C on the sense DNA strand) intronic transition, resulting in a cryptic splice acceptor and premature termination of a truncated nonfunctioning protein. *CYP3A5*\*1>*6 in rs10264272 corresponds to a G>A (C>T on the sense DNA strand) variant which causes mis-splicing and premature termination. Finally, *CYP3A5* *1>*7 in rs41303343 corresponds to a single nucleotide duplication (A>AA on the sense DNA strand), leading to a nonfunctional protein[31]. These alleles are recommended for testing due to their known effect on CYP3A5 function[29,31]. Other *CYP3A5* alleles (the *8 and *9 alleles are the only other alleles currently defined in PharmVar[51]) that may affect protein expression or function but are not associated with sufficient clinical evidence or clear treatment recommendations were not in scope of TacroType at this time. For example, the rs55817950 (*8) and rs28383479 (*9) alleles for *CYP3A5* may result in partial loss of function but their specific impact on tacrolimus metabolism has not been fully evaluated and these alleles are not associated with specific tacrolimus dosing recommendations[29,31]. Moreover, the *8 and *9 alleles are rare: *8 is reported at a frequency of 0.000024, while *9 is reported at 0.0000031[52]. *CYP3A5* *1, *3, *6 and *7 alleles, by contrast, are estimated to be present in 99-100% of various ethnic populations[53].

This study introduces TacroType™, a laboratory-developed test (LDT) for genotyping *CYP3A5* conducted at One Lambda Laboratories. The TacroType LDT combines six-plex qPCR technology with a new data analysis algorithm designed to support genotyping across a broad range of DNA concentrations.

Here we discuss the approach to multiplex genotyping assay data analysis and describe the results of our analytical verification study using a variety of well-characterized DNA and primary human samples.

## 2. Results and Discussion

### Ethnicity based analysis of CYP3A5 genotypes impacting metabolism of tacrolimus

Open sources of human genetic information including *CYP3A5* sequencing data were used to assess ethnic-specific differences affecting tacrolimus metabolism and to further evaluate the feasibility of accurately selecting treatment parameters based on ethnicity.

The current labeling for Prograf (tacrolimus) capsules, oral suspension or injections approved by the Food and Drug Administration (FDA) for use in transplant patients recommends a higher dose for patients of African descent[54]. Based on initial clinical tests performed for tacrolimus, a significant difference in pharmacokinetic data was detected between African American and Caucasian patients[41,55,56]. Subsequently, this difference was attributed to different frequencies of specific variants in the *CYP3A5* gene highly prevalent among Caucasians and less frequent in individuals of African descent[37,42,43]. To test the hypothesis that ethnicity accurately reflects genetic differences responsible for tacrolimus metabolizer status, we evaluated *CYP3A5* genotype in 2504 individuals well characterized within the 1000 Genomes Project Consortium Database. We found that nearly every ethnic group is represented by a mixture of genotypes and cannot be considered fully uniform with respect to tacrolimus metabolizer status, and that a drug dosing approach based on ethnicity alone may have a negative impact on a significant fraction of patients (**Table 1**). For example, though the general assumption of Prograf labeling is a high prevalence of rapid tacrolimus metabolizers among patients of African descent, 1000 Genome Project Consortium data demonstrate that, depending on the subpopulation, up to 30% of African-ancestry individuals may have a complete loss of CYP3A5 function (**Table 1**), a phenotype for which CPIC recommends standard tacrolimus dosing[29]. As a result, increasing starting dose based on ethnicity alone may result in supratherapeutic blood tacrolimus levels and a risk for acute kidney injury in patients of African descent. In contrast, identifying *CYP3A5* genotype prior to transplant can predict tacrolimus metabolizer status with significantly higher reliability; in a recent implementation of *CYP3A5* *3, *6 and *7 genotyping at the Indiana University School of Medicine, for example, only 66% of African Americans required a higher dose of tacrolimus[57].

**Table 1.** Percentage of tacrolimus rapid, intermediate and poor metabolizers in different human subpopulations. Metabolizer status was defined based on combined analysis of 3 common well characterized SNPs associated with loss of CYP3A5 activity.

| Population | Sub-Population | Rapid Metabolizers (%) | Intermediate Metabolizers (%) | Poor Metabolizers (%) |
| --- | --- | --- | --- | --- |
| African | African Ancestry in Southwest USA | 24.6% | 54.1% | 21.3% |
| African | African Caribbean in Barbados | 30.2% | 44.8% | 25.0% |
| African | Yoruba in Ibadan, Nigeria | 31.5% | 46.3% | 22.2% |
| African | Esan in Nigeria | 42.4% | 48.5% | 9.1% |
| African | Luhya in Webuye, Kenya | 28.3% | 47.5% | 24.2% |
| African | Gambian in Western Division, The Gambia | 24.8% | 45.1% | 30.1% |
| African | Mende in Sierra Leone | 34.1% | 49.4% | 16.5% |
| European | Finnish in Finland | 1.0% | 10.1% | 88.9% |
| European | Utah Residents with Northern and Western European Ancestry | 0.0% | 8.1% | 91.9% |
| European | Iberian Populations in Spain | 0.9% | 11.2% | 87.9% |
| European | Toscani in Italy | 0.0% | 9.4% | 90.7% |
| European | British in England and Scotland | 0.0% | 11.0% | 89.0% |
| South Asian | Bengali in Bangladesh | 14.0% | 45.4% | 40.7% |
| South Asian | Gujarati Indian in Houston, Texas | 6.8% | 41.8% | 51.5% |
| South Asian | Indian Telugu in UK | 10.8% | 43.1% | 46.1% |
| South Asian | Punjabi in Lahore, Pakistan | 16.7% | 39.6% | 43.8% |
| South Asian | Sri Lankan Tamil in the UK | 12.8% | 42.2% | 45.1% |
| East Asian | Chinese Dai in Xishuangbanna, China | 9.7% | 43.0% | 47.3% |
| East Asian | Kinh in Ho Chi Minh City, Vietnam | 7.1% | 43.4% | 49.5% |
| East Asian | Japanese in Tokyo, Japan | 3.9% | 43.3% | 52.9% |
| East Asian | Han Chinese in Beijing, China | 11.7% | 38.8% | 49.5% |
| East Asian | Southern Han Chinese, China | 7.6% | 39.1% | 53.3% |
| American | Colombian in Medellin, Colombia | 4.3% | 26.6% | 69.2% |
| American | Mexican Ancestry in Los Angeles, CA | 3.1% | 35.9% | 60.9% |
| American | Peruvian in Lima, Peru | 2.4% | 16.5% | 81.2% |
| American | Puerto Rican in Puerto Rico | 6.7% | 29.8% | 63.5% |

In addition to supratherapeutic dosing for African Americans with a poor metabolizer phenotype, standard tacrolimus dosing may under-dose patients with a rapid metabolizer phenotype. To date, there is a wide variance in patient outcomes post organ transplant. The higher incidence of adverse outcomes in African American patients [58], may be a result from lower median blood tacrolimus levels, frequently found in patients of African ancestry[42,43], and amongst African American patients, rapid *CYP3A5* metabolizers had higher first year Medicare expenses than intermediate or poor *CYP3A5* metabolizers[37]. *CYP3A5*-guided tacrolimus dosing may therefore support improved outcomes for transplant patients in impacted demographic groups.

The population data presented in **Table 1** highlights the potential utility of *CYP3A5* genotyping for patients of various ethnic backgrounds. Although current Prograf labeling does not address the presence of rapid and intermediate metabolizers in populations other than African descent, our analysis of 1000 genomes data suggests that rapid metabolizers may be present in 4 – 16% of Asian and up to 6.5% of Hispanic or Latino origins (**Table 1**). Thus, the availability of a reliable *CYP3A5* genotyping test may be beneficial for transplant patients in ethnically diverse populations.

### Development of multiplex real time PCR assay for CYP3A5 genotyping

A TaqMan-based real time PCR assay was developed for three biallelic SNPs (rs776746, rs10264272 and rs41303343) which are recommended for tacrolimus metabolism prediction by CPIC[29]. Assays were mapped and designed using a customized version of the Persephone genome browser[59].

The assay optimization process was aimed at achieving the following goals: genotyping for all 3 SNPs in a single TaqMan reaction with a sufficient level of accuracy and precision; minimizing the number of retests by achieving assay performance sufficiently robust to generate acceptable results for samples of various quality and quantity; and minimizing the impact of possible human error by introducing a simple and user-friendly workflow and data analysis process.

Primers were optimized for comparable performance efficiency in a multiplex reaction mixture to minimize the impact of competition and the risk of PCR-related artifacts. Allele-specific probes for each SNP were optimized for robust recognition of their target allele. Fluorescent labels for each probe were chosen to occupy separate excitation-emission filter combinations available on the QuantStudio 5 Dx instrument, enabling *CYP3A5* genotyping with a single six-plex reaction (**Figure 1**). A final multiplex combination was selected and optimized to maximize the signal to noise ratio for all expected genotypes.

**Figure 1.**
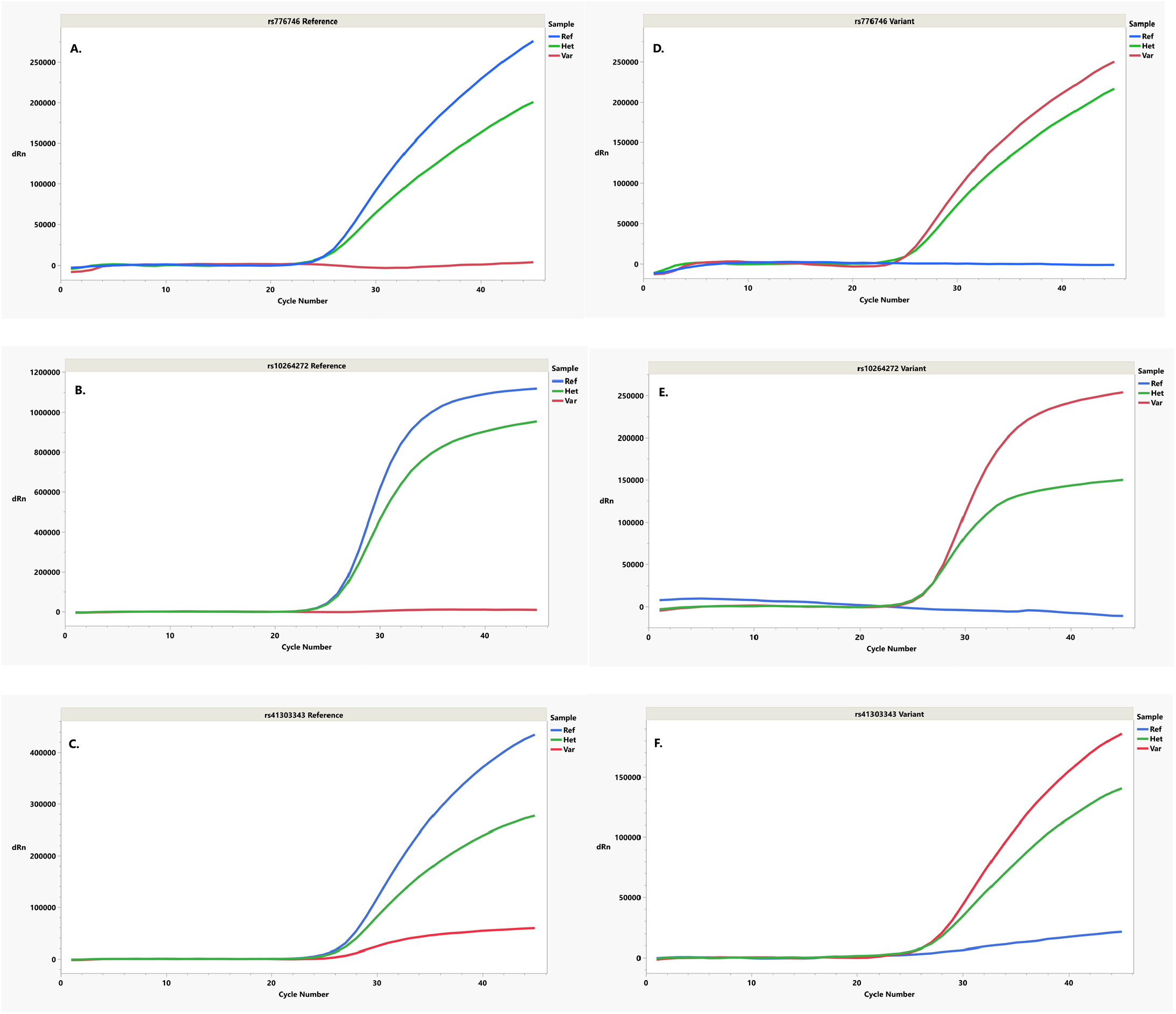
Example six-plex qPCR reaction targeting *CYP3A5* *3,*6 and *7 reference (“Ref”), heterozygote (“Het”) and variant (“Var”) groups. Representative qPCR reactions are shown for reactions targeting *CYP3A5* *3 (A,D), *6 (B,E) and *7 (C,F) reference and variant alleles.

### Genotyping data analysis algorithm

A new data analysis algorithm was developed for TacroType to reduce the impact of variability in DNA concentration across samples and to provide direct genotype output from a six-plex PCR reaction.

Although cycle of quantification (Cq value) is a reflection of DNA concentration in a qPCR reaction, amplification efficiency affects the relationship between Cq value and DNA template amount, and variable efficiency of qPCR assays may result in an inaccurate assessment of copy number with either relative or absolute quantification[60]. For multiplex reactions, competition and even small variabilities in reaction efficiencies amongst target amplicons add a layer of complexity in achieving robust and reproducible performance. Though this genotyping test was not developed for DNA quantification, DNA input levels and reaction efficiency variabilities would unavoidably impact results if a standard genotyping algorithm was used. Available TaqMan-based genotyping assays analyze end-point fluorescence results for multiple samples and evaluate data by cluster analysis of multiple reactions performed on a single plate. Normally, only fluorescence values from the first and last cycles of PCR are analyzed by genotyping algorithms[61], real time qPCR data are not used for this analysis, and as result, differences in DNA concentration and/or DNA quality within a single test may negatively impact clustering analysis and lead to inconclusive or even false calls. If this type of analysis is used, accurate results can only be achieved under standardized conditions, including a narrow range of DNA input per reaction and restrictive requirements for DNA quality.

The TacroType analysis algorithm uses real time PCR data instead of endpoint fluorescence analysis. Two types of qPCR data are exported from each test: Cq values generated with fixed thresholds; and amplification results, showing fluorescence value for each qPCR cycle. During analysis, the median of the three lowest Cq values per sample are used to calculate a sample-specific Cq. The sample-specific Cq is used as a representation of the median amplification level and efficiency for amplicons at the *3, *6 and *7 loci and accounts for possible mixtures of homozygous and heterozygous calls. The fluorescence intensity for each channel is assessed at a fixed number of cycles from the sample-specific Cq, thus standardizing results amongst different samples. This fixed number of cycles was selected to achieve an optimal signal to noise ratio for all fluorescent channels, for a variety of genotypes. This method was expected to allow significantly expanding the range of DNA inputs and quality without negatively impacting the accuracy and precision of the genotyping assay.

While standardized fluorescence values can be analyzed by clustering algorithms or a method relying on fixed cutoff values for each fluorophore, a combination of both methods can also be used. A locked version of a data analysis tool based on our proprietary genotyping algorithm was tested together with the TacroType assay within an analytical validation study to evaluate the accuracy and precision of new laboratory developed test.

We expected that implementation of a six-plex assay with six distinct fluorophores and a new analysis algorithm would reduce variability associated with three known sources: biological, protocol and technical, highlighted by MIQE guidelines[62]. Probe-based discrimination assays using end-point fluorescence for genotyping purposes require strict normalization of input DNA, since the fluorescence level at any given cycle of a qPCR reaction depends on the number of cycles elapsed from the start of amplification. Maintaining strict DNA input for large numbers of clinical samples is not only laborious but also challenging due to potential differences in DNA quality (biological variability) as well as possible human error (technical variability). We implemented analysis of fluorescence at a fixed number of cycles following the sample specific Cq value, reducing the impact of potential errors generated in the process of measuring and normalizing samples before performing qPCR. Furthermore, since multiplexing is performed with a separate fluorophore for each individual assay tested, protocol variability is reduced, since there is a reduced risk of operators mixing primer/probe mixes for separate assays. Finally, since the entire workflow involves fewer reaction mixes, and therefore fewer pipetting steps, implementation of a single multiplex reaction mix reduces technical variability.

An analytical validation study was designed to test these assumptions and evaluate the accuracy and precision of the complete laboratory-developed test (LDT) process from sample preparation through the genotyping report generated by the data analysis algorithm.

Full analytical validation of the LDT process was performed following CLSI guidelines[63].

### Analytical accuracy and precision of TacroType evaluated with well-characterized DNA samples

A panel of thirty-two well-characterized DNA samples was utilized (**Table 13**) to perform the initial evaluation of accuracy, reproducibility and repeatability for TacroType. The panel included multiple samples from each possible *CYP3A5* genotype (**Table 2**). All samples were dispensed in triplicate per 96 well plate using 25 ng of total DNA per reaction. Genotyping concordance in each test was analyzed comparing each pair of alleles reported by our data analysis algorithm to known typing of each reference DNA.

**Table 2.** Summary of quantity of samples utilized from each genotype for verification studies.

| Source | *1/*1 | *1/*3 | *1/*6 | *1/*7 | *3/*6 | *3/*7 | *6/*7 | *3/*3 | *6/*6 | *7/*7 |
| --- | --- | --- | --- | --- | --- | --- | --- | --- | --- | --- |
| Coriell | 2 | 0 | 0 | 0 | 2 | 2 | 3 | 2 | 2 | 2 |
| One<br>Lambda | 3 | 2 | 3 | 2 | 2 | 2 | 1 | 2 | 0 | 0 |

In initial accuracy testing, TacroType generated results 100% concordant with reference genotypes for tested DNAs (lower bound of 95% confidence interval for allelic call concordance was 98.4%).

To test reproducibility amongst multiple lots of reagents, three different lots were tested in duplicate using the same well-characterized DNA samples. All 3 lots generated results concordant to reference typing of the samples with 100% concordance amongst lots of reagents, providing overall 100% accuracy with a lower bound 95% confidence interval of 97.7% per lot.

For initial precision evaluation, the same panel of thirty-two well-characterized DNA samples dispensed in triplicate was tested by two operators twice a day (morning and afternoon) using two different instrument types through 3 non-consecutive days (**Table 3**). The reproducibility and repeatability study included 2304 individual qPCR reactions with 99.8% of reactions generating genotyping results in our data analysis algorithm. Four cases of sample amplification failure did not correlate with any specific confounder such as operator, instrument or time of the day, and were attributed to random technical errors during qPCR reaction setup. The remaining 2300 qPCR reactions provided reportable results, which were 100% concordant to reference typing. A summary of the precision study and confidence intervals for each evaluated variable is presented in **Table 3**.

**Table 3.** Summary of precision studies performed using well-characterized DNA samples.

| Factor Tested | Groups Tested | Plates/Group | Samples-Group | Minimal reported genotypes per group, % | Concordance, % | Lower Bound of 95% Confidence Interval<sup>1</sup> |
| --- | --- | --- | --- | --- | --- | --- |
| Time of Day | AM, PM | 12 / AM or PM | 1152 | 99.8% | 100% | 99.8% |
| Day | Six days | 4 / day | 384 | 99.7% | 100% | 99.5% |
| Operator | 1, 2 | 12 / operator | 1152 | 99.8% | 100% | 99.8% |
| Instrument | 1, 2, 3, 4 | 6 / instrument | 576 | 99.6% | 100% | 99.6% |

<sup>1</sup>One sided low bound 95% confidence interval is calculated by exact binomial test for each evaluated category

### Measuring range using well-characterized DNA samples

Analytical range was initially estimated using well-characterized DNA samples enabling testing heterozygous and homozygous samples for each allele at a range of concentrations from 0.5-1600 ng (**Table 4, Figure 2**). For each DNA input, each sample was tested using two separate lots of primer/probe mix in triplicate. The total number of reactions per DNA concentration was 72 (12 unique samples, 2 lots of reagents, 3 replicates), resulting in 144 allelic calls per each DNA input. In a total of 576 qPCR reactions (72 tests for 8 DNA concentrations) genotyping results were generated for 99.7% of reactions. Two cases of amplification failure were not associated with a specific DNA input or reagent lot and were attributed to random technical errors. For all reported genotypes, 100% concordance to the reference typing was observed with a lower bound 95% confidence interval exceeding 97.9% calculated per DNA concentration using the exact binomial test.

**Table 4.**
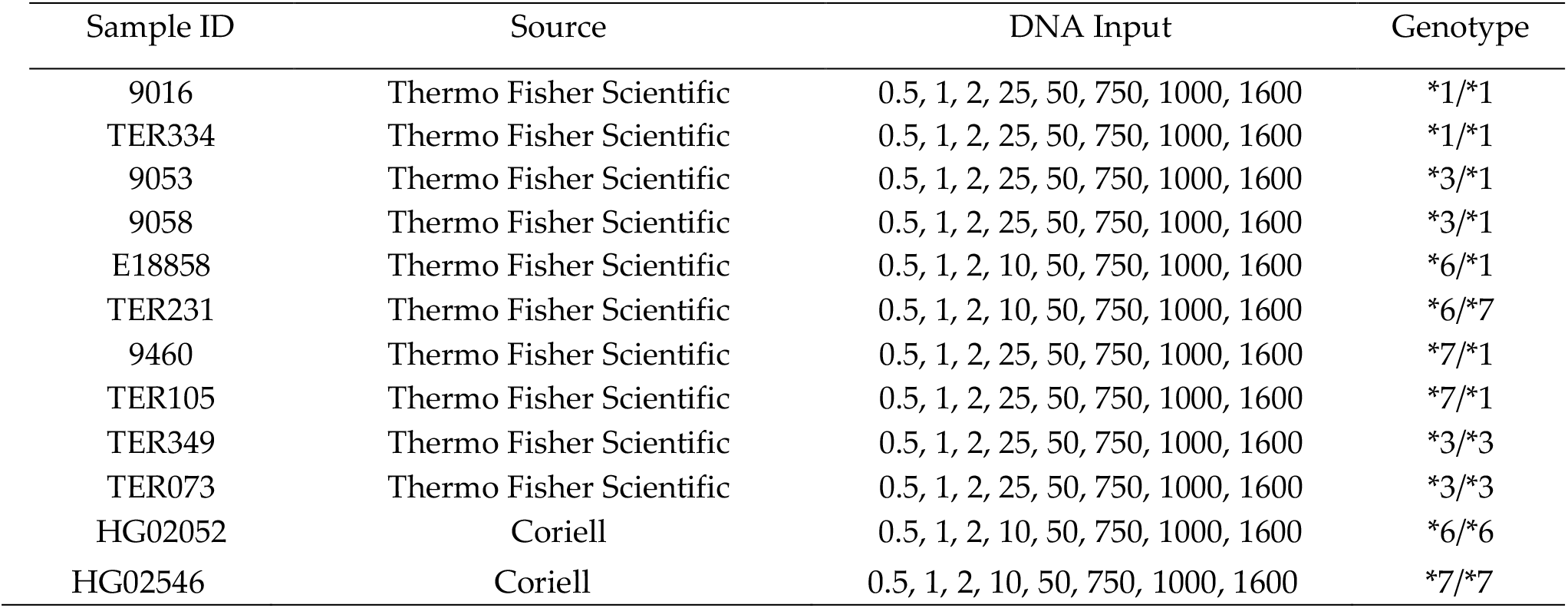
Summary of DNA sample, source, input and genotype information for analytical range studies.

**Figure 2.**
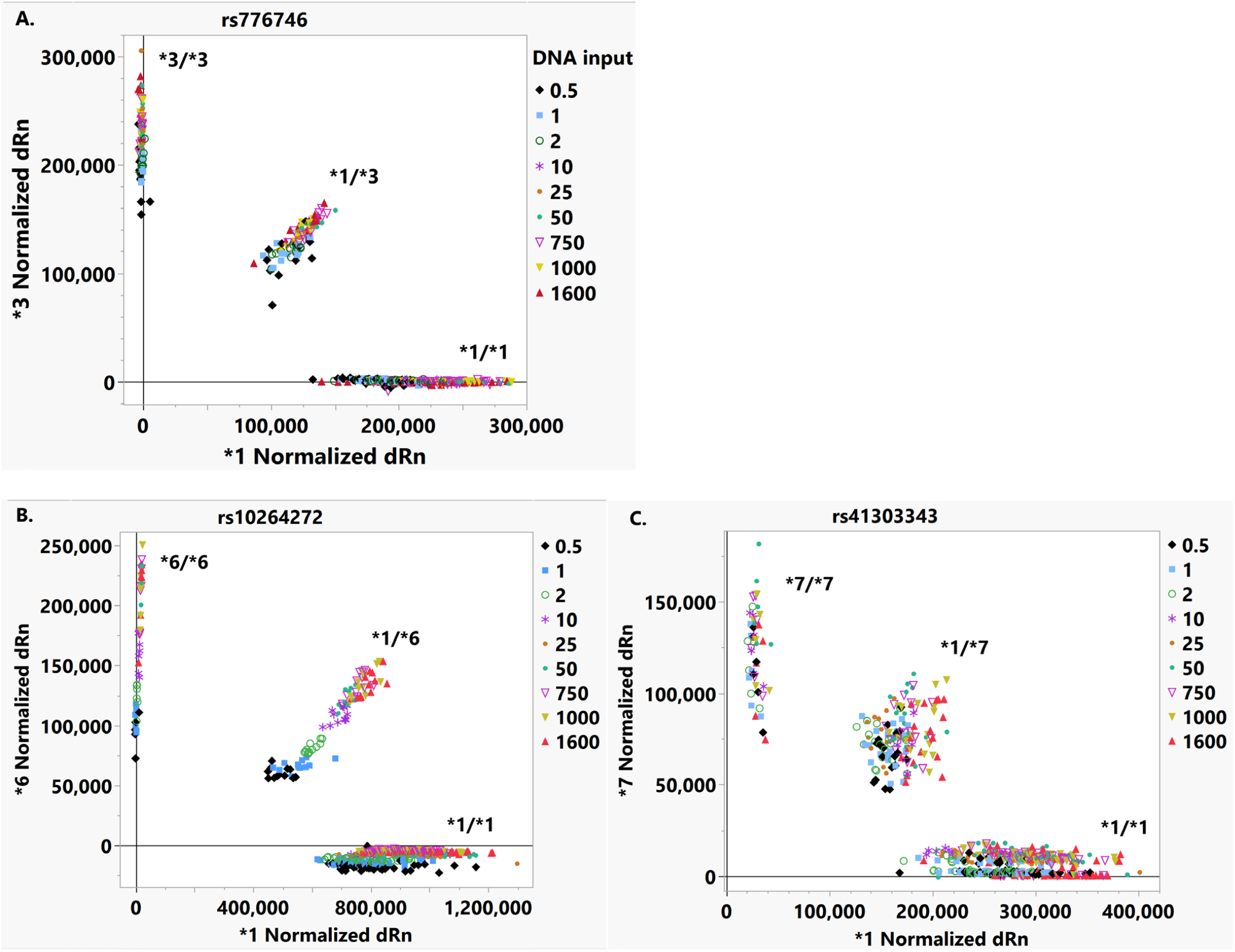
TacroType performance across a range of sample inputs. Cluster plots are shown for *CYP3A5* *3 (A), *6 (B) and *7 (C) alleles. Samples clustered into corresponding genotyping groups when DNA inputs ranging from 0.5 ng to 1600 ng were utilized.

Figure 2. shows the results of clustering analysis for a measuring range study illustrating how our data analysis approach allows standardization of results from TaqMan reactions testing DNA input spanning over 3 logarithms range, with fluorescence values still generating distinct clusters per each genotype.

Thus, the DNA input range suitable for TacroType was established as 0.5 – 1600 ng per reaction. Concentrations below 0.5 ng per reaction will require re-extraction, and are not recommended for testing due to the risk of allelic imbalance resulting from random sampling variability when insufficient DNA input is used for genotyping purposes.

### Sample Preparation Study

After confirming the performance of TacroType within a defined DNA input range using high quality well-characterized DNA samples, we evaluated the accuracy and precision of TacroType using a variety of primary human samples.

Accuracy studies were performed with 40 EDTA blood samples obtained from DLS. All blood samples generated concordant genotyping results when compared to a commercially available genotyping test (lower bound of 95% CI = 96.3%).

We expanded the scope of our study to a larger variety of sample collection methods. The following sample types were evaluated: whole blood collected using ethylenediaminetetraacetic acid (EDTA), acid citrate dextrose (ACD) and PAXgene blood collection tubes; and buccal swabs collected using HydraFlock swabs (Puritan). Blood and buccal samples were stored at ambient temperature for up to 72 hours prior to extraction without significant impact on TacroType results. Blood frozen at -20°C and -80°C was also tested to verify conditions suitable for extended sample storage.

Though the primary method selected for the LDT process was automated DNA extraction using the EZ1&2 DNA Tissue Kit (Qiagen), secondary methods were also evaluated to understand whether TacroType is limited by specific sample quality requirements. The range of DNA quantity and quality for each sample type and DNA extraction method is shown in **Table 5** (236 DNA extractions evaluated by nanodrop spectrometer) and **Table 6** (110 DNA extractions evaluated by Qubit fluorometer). In a total of 346 DNA samples extracted by 6 methods, 100% of samples contained DNA concentrations within the range acceptable for use with TacroType without additional normalization. The study included samples from 7 different sample collection events including a total of 160 healthy donors (**Table 7**).

**Table 5.**
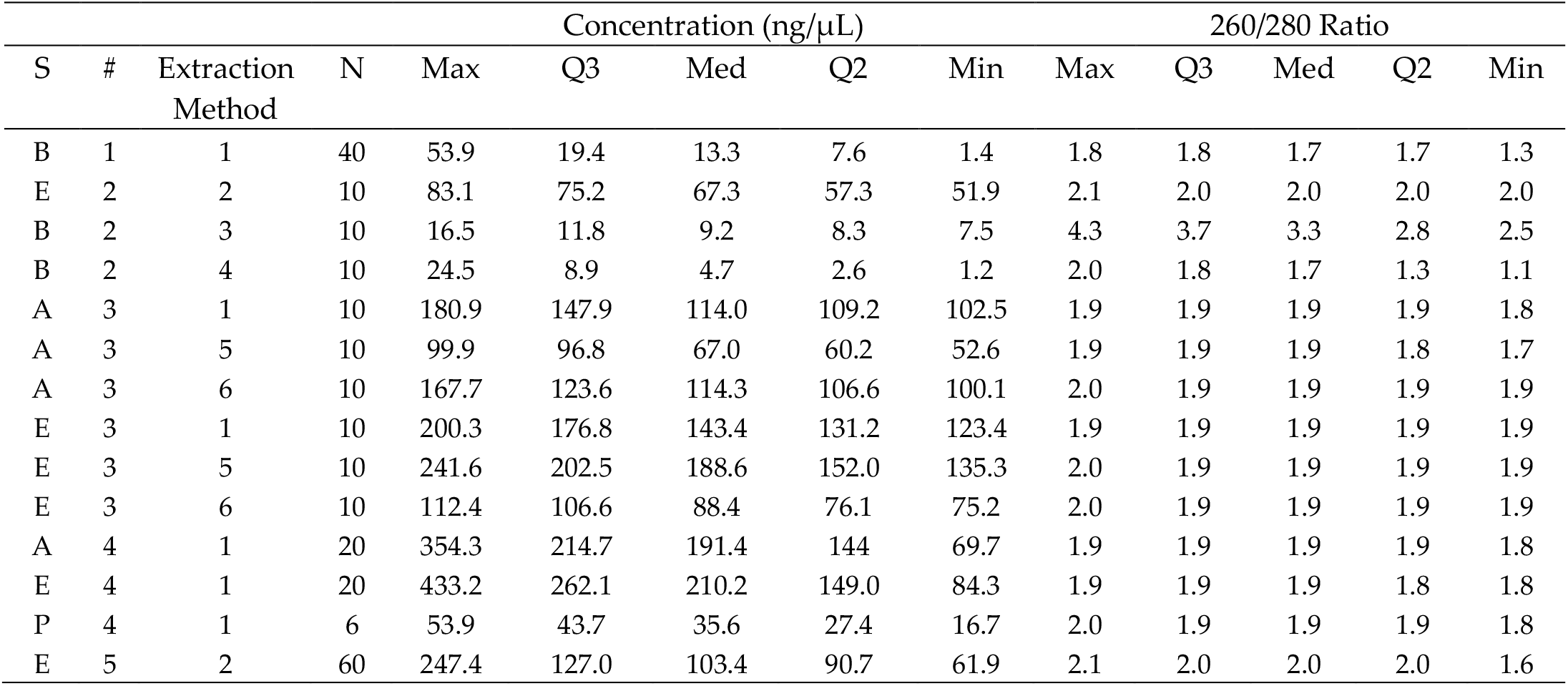
Summary of DNA concentration and purity measurements following blood or buccal swab extraction using nanodrop spectrometer. Q3, top quartile. Q2, bottom quartile. S, source. B, buccal swab; E, blood EDTA collection tubes; P, PAXgene Blood RNA collection tubes; A, Blood ACD collection tubes. #, group number. Extraction methods are as follows: 1-Dynabeads; 2, EZ1/2 Virus Mini Kit; 3, EZ1/2 DNA Tissue Kit; 4, QIAamp DNA mini kit; 5, Maxwell RSC Whole Blood DNA Kit; 6, QIAamp DNA Blood midi kit.

**Table 6.** Summary of DNA concentration following blood or buccal swab DNA extraction. DNA concentration was measured using Qubit. Q3, 75% percentile. Q2, 25% percentile.

| Source | Extraction Method | N | Max | Concentration (ng/ μL) |  |  |  |
| --- | --- | --- | --- | --- | --- | --- | --- |
|  |  |  |  | Q3 | Med | Q2 | Min |
| Buccal Swabs | EZ1/2 DNA Tissue Kit | 40 | 29.7 | 13.9 | 9.0 | 6.3 | 1.1 |
| Buccal Swabs | QIAamp DNA Mini Kit | 10 | 35.5 | 20.9 | 15.7 | 8.9 | 5.9 |
| EDTA Blood | EZ1/2 DNA Tissue Kit | 40 | 65.0 | 47.6 | 42.6 | 35.3 | 23.1 |
| EDTA Blood | QIAamp DNA Blood Mini Kit | 20 | 70.0 | 43.5 | 23.4 | 16.1 | 7.4 |

**Table 7.** Summary of results from sample preparation studies during assay verification. RT, room temperature.

| Specimen Type | Storage Conditions | Extraction Method | Samples | Reportable Results (%) | Concordance<sup>1</sup> (%) |
| --- | --- | --- | --- | --- | --- |
| Sample Set 1: total concordance 100% with lower CI 96.3 |  |  |  |  |  |
| Buccal Swabs | RT 24 hrs | Dynabeads | 20 | 100 | 100 |
| Buccal Swabs | RT 72 hrs | Dynabeads | 20 | 100 | 100 |
| Sample Set 2: total concordance 100% with lower CI 95.1 |  |  |  |  |  |
| Buccal Swabs | RT 48 hrs | QIAamp DNA Mini Kit | 10 | 100 | 100 |
| Buccal Swabs | RT 48 hrs | EZ1/2 DNA Tissue Kit | 10 | 100 | 100 |
| Blood (EDTA) | -20°C | EZ1/2 Virus Mini Kit | 10 | 100 | 100 |
| Sample Set 3: total concordance 100% with lower CI 97.5 |  |  |  |  |  |
| Blood (ACD) | 4°C 48 hrs | Dynabeads | 10 | 100 | 100 |
| Blood (ACD) | 4°C 48 hrs | QIAamp DNA Blood Midi Kit | 10 | 100 | 100 |
| Blood (ACD) | 4°C 48 hrs | Maxwell | 10 | 100 | 100 |
| Blood (EDTA) | 4°C 48 hrs | Dynabeads | 10 | 100 | 100 |
| Blood (EDTA) | 4°C 48 hrs | QIAamp DNA Blood Midi Kit | 10 | 100 | 100 |
| Blood (EDTA) | 4°C 48 hrs | Maxwell | 10 | 100 | 100 |
| Sample Set 4<sup>2</sup>: total concordance 100% with CI 96.7 |  |  |  |  |  |
| Blood (ACD) | 4°C 24 hrs | Dynabeads | 20 | 100 | 100 |
| Blood (EDTA) | 4°C 24 hrs | Dynabeads | 20 | 100 | 100 |
| Blood (PAXgene) | 4°C 24 hrs | Dynabeads | 6 | 100 | 100 |
| Intra-condition concordance |  |  |  |  | 100 |
| Sample Set 5: total concordance 100% with CI 97.5 |  |  |  |  |  |
| Blood (EDTA) | 4°C 24 hrs | EZ1/2 Virus Mini Kit | 20 | 100 | 100 |
| Blood (EDTA) | 4°C 48 hrs | EZ1/2 Virus Mini Kit | 20 | 100 | 100 |
| Blood (EDTA) | -80°C | EZ1/2 Virus Mini Kit | 20 | 100 | 100 |
| Sample Set 6: total concordance 100% with CI 97.0 |  |  |  |  |  |
| Buccal Swabs | RT <72 hrs | QIAamp DNA Mini Kit | 10 | 100 | 100 |
| Buccal Swabs | RT <72 hrs | EZ1/2 DNA Tissue Kit | 10 | 100 | 100 |
| Buccal Swabs | RT < 72 hrs | EZ1/2 DNA Tissue Kit | 30 | 100 | 100 |
| Sample Set 7: total concordance 100% with CI 97.5 |  |  |  |  |  |
| Blood (EDTA) | 4°C < 15d | QIAamp DNA Blood Mini Kit | 20 | 100 | 100 |
| Blood (EDTA) | 4°C < 15d | EZ1/2 DNA Tissue kit | 20 | 100 | 100 |
| Blood (EDTA) | 4°C < 15d | EZ1/2 DNA Tissue Kit | 20 | 100 | 100 |

<sup>1</sup>Concordance is defined as agreement with genotyping results obtained using TaqMan assays, excluding the combination of sample failures and technical failures leading to no result. DNA input was 3 µL of undiluted genomic DNA (sample set 1), 4 µL of undiluted genomic DNA (sample set 2), 25 ng (sample set 3), 19 ng (sample set 4), 2 µL of undiluted genomic DNA (sample set 5) and 4 µL of undiluted genomic DNA (sample set 6-7).
<sup>2</sup>Sample set 4 included 20 separate blood donors. We extracted ACD and EDTA blood tubes for all 20 donors, and PAXgene blood tubes for 6 donors.

The sample preparation study demonstrated that TacroType is suitable for use with DNA extracted from buccal swabs stored for up to 72 hours at ambient temperature post-collection. Buccal swabs extracted with three separate DNA extraction methods generated acceptable results (**Table 7**). Both ACD and EDTA blood samples were suitable for use with TacroType, for a range of storage conditions indicated in **Table 7**.

### Measuring limit for primary human samples

DNA extracted from a variety of human samples by multiple sample preparation methods demonstrated sample concentrations within the TacroType measuring range for 100% of tested samples (**Table 5-6**). Nevertheless, we performed additional analytical tests to confirm that the lower limit of the acceptable DNA input range for primary human samples is comparable to results obtained from initially tested well-characterized DNA (**Figure 2**) and primary human samples prepared by selected methods (**Table 7**).

A minimal DNA input of 0.5 ng was tested for DNA extracted from blood EDTA and buccal swab samples (**Table 8**). Each of 10 buccal swabs or 20 blood samples collected in EDTA tubes was tested in duplicate using two separate instruments. At the 0.5 ng DNA per reaction, concordance was 100% with a lower bound of the 95% confidence interval of 96.3% for buccal swabs and 98.1% for EDTA blood samples.

**Table 8.** Summary of measuring limit studies performed using primary human samples.

| Sample Type | DNA Input | Total qPCR reactions<br>in the study | Concordance %<br>(95% CI) |
| --- | --- | --- | --- |
| Blood (EDTA) | 0.5 ng | 80 | 100% (98.1%) |
| Buccal Swabs | 0.5 ng | 40 | 100% (96.3%) |

### Analytical precision: Repeatability and Reproducibility in primary human samples

Repeatability and reproducibility studies were performed for both blood and buccal swabs.

For blood studies, two operators tested DNA extracted from the blood of 5 different donors using 2 different qPCR instruments, and samples were tested in duplicates for all tested conditions. This study assessed the effects of time of day (5 samples in duplicate on two separate instruments, AM or PM), day of test (5 samples in duplicate on two separate instruments, 3 consecutive days), and technician (5 samples in duplicate on two separate instruments, 2 technicians) on assay performance. This precision study showed 100% genotyping reproducibility in blood samples for tested conditions with a lower bound confidence interval for concordance of 99.2%. Sample counts and results are summarized in **Table 9**.

**Table 9.** Summary of factors tested for assessment of repeatability and reproducibility for blood samples collected in EDTA tubes.

| Factor Tested | Groups Tested | Plates/Group | Total qPCR reactions in the study | Concordance %<br>(95% CI) |
| --- | --- | --- | --- | --- |
| Time of Day | AM, PM | 2/ AM or PM | 40 | 100% (96.3%) |
| Day | Three consecutive days | 2 / day | 60 | 100% (97.5%) |
| Operator | 1, 2 | 2 / operator | 40 | 100% (96.3%) |
| Instrument<sup>1</sup> | 1, 2 | 4 / instrument | 180 | 100% (99.2%) |

<sup>1</sup>Instrument studies included 140 qPCR reactions from preceding factors, plus an additional 40 qPCR reactions in a dedicated instrument test.

For buccal swab samples two operators tested DNA extracted from buccal swabs from 5 different donors using 2 different qPCR instruments, and samples were tested in duplicates for all tested conditions. The effects of time of day (5 samples in duplicate on two separate instruments, AM or PM), day of test (5 samples in duplicate on two separate instruments, 3 consecutive days) and technician (5 samples in duplicate on two separate instruments, 2 technicians) on assay performance were assessed. The buccal swab precision study showed 100% genotyping reproducibility in buccal swab samples for tested conditions with a lower bound confidence interval for concordance of 99.3%. Sample counts and results are summarized in **Table 10**.

**Table 10.** Summary of factors tested for assessment of repeatability and reproducibility for buccal swab samples.

| Factor Tested | Groups Tested | Plates/Group | Total qPCR reactions in the<br>study | Concordance %<br>(95% CI) |
| --- | --- | --- | --- | --- |
| Time of Day | AM, PM | 2/ AM or PM | 40 | 100% (96.3%) |
| Day | Three consecutive days | 2 / day | 60 | 100% (97.5%) |
| Operator | 1, 2 | 2 / day | 40 | 100% (96.3%) |
| Instrument<sup>1</sup> | 1, 2 | 4 / instrument | 200 | 100% (99.3%) |

<sup>1</sup>Instrument studies included 140 qPCR reactions from preceding studies, plus 60 qPCR reactions in dedicated instrument test.

### Interfering Substances

DNA amplification-based methods may be significantly impacted by sample components if they carry over through the sample preparation process. DNA extraction reagents may also contaminate human samples used for laboratory testing. For human genotyping tests, the main expected impact of interfering substances is inhibition of amplification, resulting in lower signals or amplification failures.

To evaluate the direct impact of known interferants on DNA samples extracted from whole blood, DNA extracted from EDTA blood collection tubes was spiked with heparin or hemin chloride to assess their inhibitory effect at different concentrations. A range of concentrations of each inhibitor on a single sample was tested to initially determine the concentrations of each contaminant expected to result in inhibition of amplification. Concentrations greater than 25 ng/µL resulted in complete inhibition of PCR amplification.

Based on initial results with a range of interferant concentrations, two conditions for each interferant were assessed: firstly, a condition in which 5 ng of DNA was tested in the presence of a concentration of interferant expected to result in inhibition; and secondly, a five-fold dilution of the DNA and interferent in the first condition which is expected to be below the concentration of interferant that results in inhibition of amplification while still maintaining DNA input acceptable for TacroType (**Table 11**). Five-fold dilution rescued qPCR performance when DNA was spiked with either 50 ng/µL heparin or 100 ng/µL hemin (**Table 11**).

**Table 11.** Summary of interfering substances studies using either EDTA blood collection tubes or buccal swab samples.

| Specimen Type | Final Inhibitor Concentration | Samples | Reportable Results (%) | Concordance (%) |
| --- | --- | --- | --- | --- |
| Blood (EDTA) | 50 ng/μL heparin | 10 | 0% | N/A |
| Blood (EDTA) | 10 ng/μL heparin | 10 | 100% | 100% |
| Blood (EDTA) | 100 ng/μL hemin | 10 | 0% | N/A |
| Blood (EDTA) | 20 ng/μL hemin | 10 | 100% | 100% |
| Buccal Swabs | Buccal Swabs | 24 | 100% | 100% |

Interfering substances studies with buccal swabs were performed by using donor samples in which donors did not follow standard buccal swab collection procedures (i.e.; no food or drink for at least 30 minutes before collection; **Table 11**). Paired samples collected by the same donors following buccal swab collection protocol were used as a control in this study.

Importantly, though the presence of some interferents could lead to amplification failure and a lack of genotyping results reported for affected samples, we did not observe any cases of inaccurate genotyping. These studies indicate a risk of retest associated with biological interference or technical variability but no detectable risk to genotyping accuracy for TacroType.

In summary, the analytical study included 3168 qPCR reactions with well-characterized reference DNA samples and 846 reactions with a variety of primary human samples prepared by different DNA extraction methods. A total of 6 failures to generate genotyping results represent 0.15% of total tests, indicating a relatively low risk of retests associated with sample quality or technical variabilities. Moreover, 100% of reported genotypes were accurate, suggesting overall accuracy of the test exceeding 99.9% (lower 95% CI for exact binomial test)

This analytical study confirms our initial assumptions that the new multiplex PCR and data analysis workflow enables reducing the impact of known variabilities associated with PCR-based measurement methods. A potential challenge in designing endpoint-based fluorescence assays is variability in concentrations and quality amongst biological samples. Standardization of DNA input is one solution to this challenge, however this involves additional pipetting steps, during which technical variability may be introduced. This approach may also increase the frequency of DNA re-extraction and the associated increase in turnaround time. For endpoint-based fluorescence tests, assessment of ΔRn values at a set number of cycles following the Cq value standardizes signal amongst a wide range of DNA inputs, reducing the need for concentration adjustments.

A genotyping process using a higher multiplex level reduces the risk of technical errors such as sample switch, since only a single Pmix is tested for each DNA sample. Moreover, the reduction in the number of Pmixes greatly mitigates protocol variability associated with plate position or time each reagent is handled across laboratories or operators. We demonstrated that robust performance of a high multiplex TaqMan assay can be achieved with consistent accuracy of genotyping results.

Biological variability introduced by differences in DNA quality between samples and by the presence of natural interferents was also evaluated with no detectable impact on genotyping accuracy of the TacroType LDT. We also confirmed that TacroType performed accurately across a wide range of DNA concentrations and quality characterized by differences in 260/280 ratios (**Table 5-7**). We expect that TacroType significantly mitigates all three sources of qPCR variability identified in MIQE 2.0 guidelines[62] and will contribute to further improvement of post-transplant care with particular importance for organ and hematopoietic transplant patients treated by tacrolimus-based medications.

Accurate assessment of the relationship between *CYP3A5* genotype and tacrolimus dosing requires testing for all three actionable *CYP3A5* alleles, particularly for populations including individuals of African descent who may have higher chance to be affected by more than one genetic variant associated with the loss of CYP3A5 function. Thus, based on 1000 genomes data, the frequency distribution of the *6 allele in different populations is 0.154 in African, 0.023 in American, 0.002 in European and 0 in Asian populations. The *7 allele was detected with a frequency of 0.118 in African, 0.003 in American and 0 in all other populations. Therefore, *CYP3A5* genotyping methods targeting only *1 and *3 alleles are significantly less reliable in predicting tacrolimus metabolism in patients of African descent.

Including *6 and *7 typing is essential for reliable *CYP3A5* genotyping in ethnically diverse populations. Review of published clinical trials data where *CYP3A5* genotype was analyzed demonstrated that only ∼5% of all trials included testing for *6 and *7 alleles (**Table 12**). The studies in which the *6 and *7 alleles were not tested included a trial in which liver donor ethnicity was not clearly stated[38] as well as a study that included approximately 40 % African Americans[39]. Similarly, the 2016 prospective clinical trial performed by Shuker and colleagues[36] did not assess the *CYP3A5* *6 or *7 alleles, despite 23/203 (11.3%) of the study population being of African descent. Amongst *CYP3A5* expressors, significantly more supratherapeutic drug concentrations were observed in the genotype-guided arm (13/28, 46.4%), as compared to the standard dose arm (3/23, 13%; p = 0.024) at day 3 following transplantation. Since apparent *CYP3A5* expressors were given twice the dose of *CYP3A5* non-expressors, the supratherapeutic genotype-guided dosing observed in the Shuker et al study may well be a result of inaccurate assessment of genotypes. Out of 661 individuals of African descent in the Ensembl database[64], we identified 304 (46%) that carried *CYP3A5* *6 and/or *CYP3A5*\*7 alleles, indicating a major risk for incorrect gene-drug associations in studies with a significant percentage of individuals of African descent. This represents a clear example illustrating the recommendation in MIQE 2.0 to “consider the biological context” of a qPCR-based test[62].

**Table 12.**
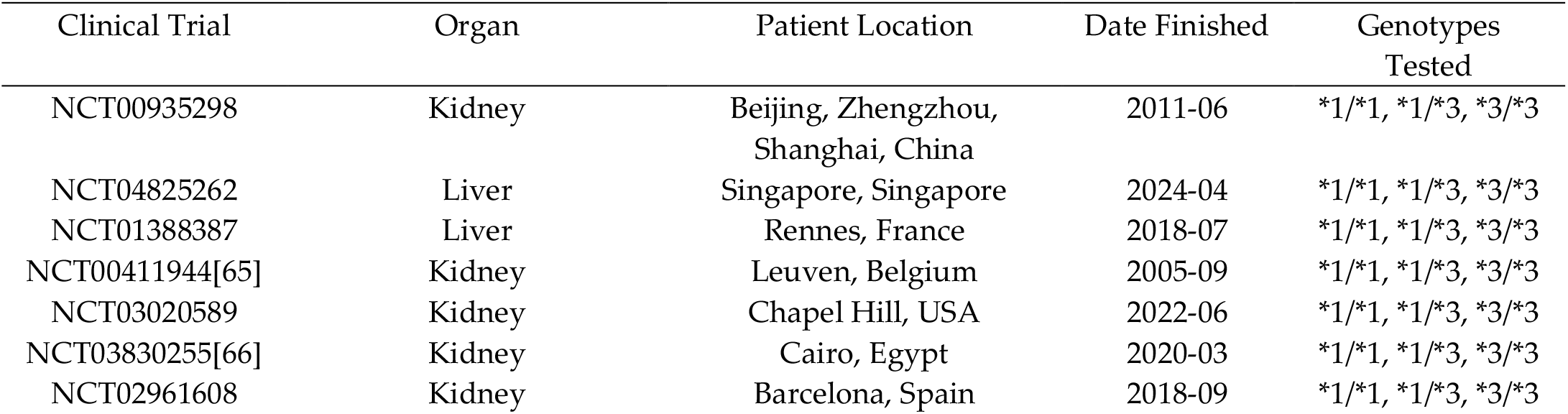
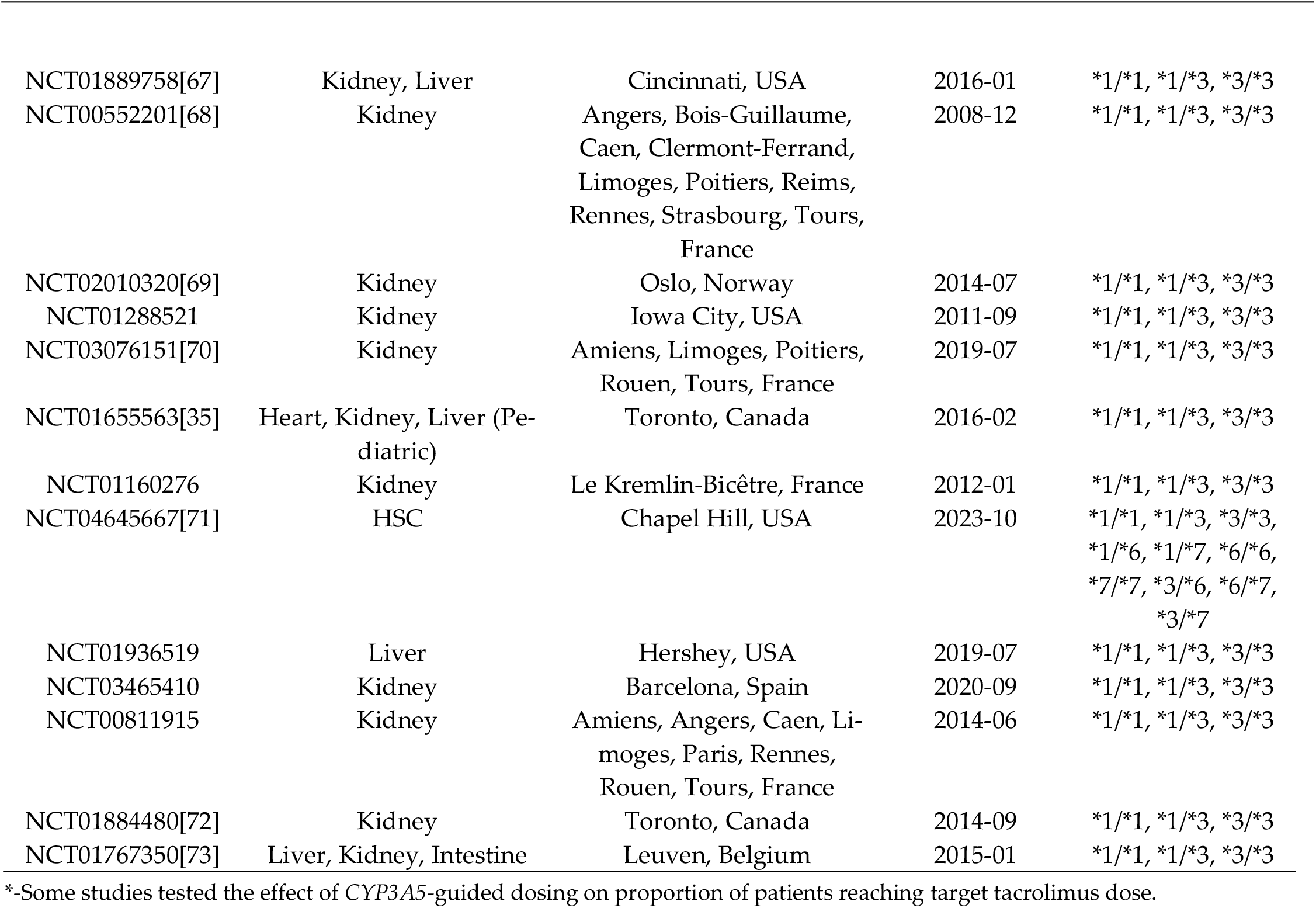
Summary of clinical trials and used to assess the relationship between *CYP3A5* and tacrolimus dosing. Clinical trials were obtained from https://clinicaltrials.gov, using the terms “*CYP3A5*” and “Tacrolimus” and filtering for completed studies. When information was not available on clinicaltrials.gov, a reference is provided. Studies in which transplant patients were not included, which were not performed due to lack of patients, or for which demographic information could not be obtained were excluded. HSC, hematopoietic stem cells.

| Clinical Trial | Organ | Patient Location | Date Finished | Genotypes Tested |
| --- | --- | --- | --- | --- |
| NCT00935298 | Kidney | Beijing, Zhengzhou, Shanghai, China | 2011-06 | *1/*1, *1/*3, *3/*3 |
| NCT04825262 | Liver | Singapore, Singapore | 2024-04 | *1/*1, *1/*3, *3/*3 |
| NCT01388387 | Liver | Rennes, France | 2018-07 | *1/*1, *1/*3, *3/*3 |
| NCT00411944[65] | Kidney | Leuven, Belgium | 2005-09 | *1/*1, *1/*3, *3/*3 |
| NCT03020589 | Kidney | Chapel Hill, USA | 2022-06 | *1/*1, *1/*3, *3/*3 |
| NCT03830255[66] | Kidney | Cairo, Egypt | 2020-03 | *1/*1, *1/*3, *3/*3 |
| NCT02961608 | Kidney | Barcelona, Spain | 2018-09 | *1/*1, *1/*3, *3/*3 |
| NCT01889758[67] | Kidney, Liver | Cincinnati, USA | 2016-01 | *1/*1, *1/*3, *3/*3 |
| NCT00552201[68] | Kidney | Angers, Bois-Guillaume, Caen, Clermont-Ferrand, Limoges, Poitiers, Reims, Rennes, Strasbourg, Tours, France | 2008-12 | *1/*1, *1/*3, *3/*3 |
| NCT02010320[69] | Kidney | Oslo, Norway | 2014-07 | *1/*1, *1/*3, *3/*3 |
| NCT01288521 | Kidney | Iowa City, USA | 2011-09 | *1/*1, *1/*3, *3/*3 |
| NCT03076151[70] | Kidney | Amiens, Limoges, Poitiers, Rouen, Tours, France | 2019-07 | *1/*1, *1/*3, *3/*3 |
| NCT01655563[35] | Heart, Kidney, Liver (Pediatric) | Toronto, Canada | 2016-02 | *1/*1, *1/*3, *3/*3 |
| NCT01160276 | Kidney | Le Kremlin-Bicêtre, France | 2012-01 | *1/*1, *1/*3, *3/*3 |
| NCT04645667[71] | HSC | Chapel Hill, USA | 2023-10 | *1/*1, *1/*3, *3/*3, *1/*6, *1/*7, *6/*6, *7/*7, *3/*6, *6/*7, *3/*7 |
| NCT01936519 | Liver | Hershey, USA | 2019-07 | *1/*1, *1/*3, *3/*3 |
| NCT03465410 | Kidney | Barcelona, Spain | 2020-09 | *1/*1, *1/*3, *3/*3 |
| NCT00811915 | Kidney | Amiens, Angers, Caen, Limoges, Paris, Rennes, Rouen, Tours, France | 2014-06 | *1/*1, *1/*3, *3/*3 |
| NCT01884480[72] | Kidney | Toronto, Canada | 2014-09 | *1/*1, *1/*3, *3/*3 |
| NCT01767350[73] | Liver, Kidney, Intestine | Leuven, Belgium | 2015-01 | *1/*1, *1/*3, *3/*3 |

\*-Some studies tested the effect of CYP3A5-guided dosing on proportion of patients reaching target tacrolimus dose.

Results of these clinical trials indicate a need for a simple genotyping solution to ensure accurate assessment of *CYP3A5* genotype. The technology implemented for TacroType provides a simple workflow enabling accurate and robust performance of a genotyping test within a laboratory environment.

TacroType is used for clinical purposes by the CLIA-certified laboratory performing the test. It has not been cleared or approved by the U.S. Food and Drug Administration or CE marked in the European Union as an in vitro diagnostic test. Clinical management of tacrolimus therapy should follow local guidelines and include therapeutic drug monitoring and other patient assessments if required.

## 3. Materials and Methods

### Analysis of the CYP3A5 gene in different ethnic groups using SNP data from the 1000 Genomes Project Consortium

To assess the ethnic distribution of *CYP3A5* expressor status based on the three most common SNPs, we utilized the Ensembl 2025 database for 2504 individual samples from the 1000 Genomes Project Consortium[74] with known genotype at the rs776746 (*3), rs10264272 (*6) and rs41303343 (*7) loci[64]. Individuals that had the allele corresponding to *1 at all three loci were defined as rapid metabolizers, individuals that were heterozygotes for *1 and either *3, *6 or *7 were counted as intermediate metabolizers, and individuals that had an allele associated with loss of function in both chromosomes (i.e; *3/*3 or *6/*7) were identified as poor metabolizers.

### DNA samples

Well-characterized DNA samples used in the study are listed in **Table 13**.

**Table 13.**
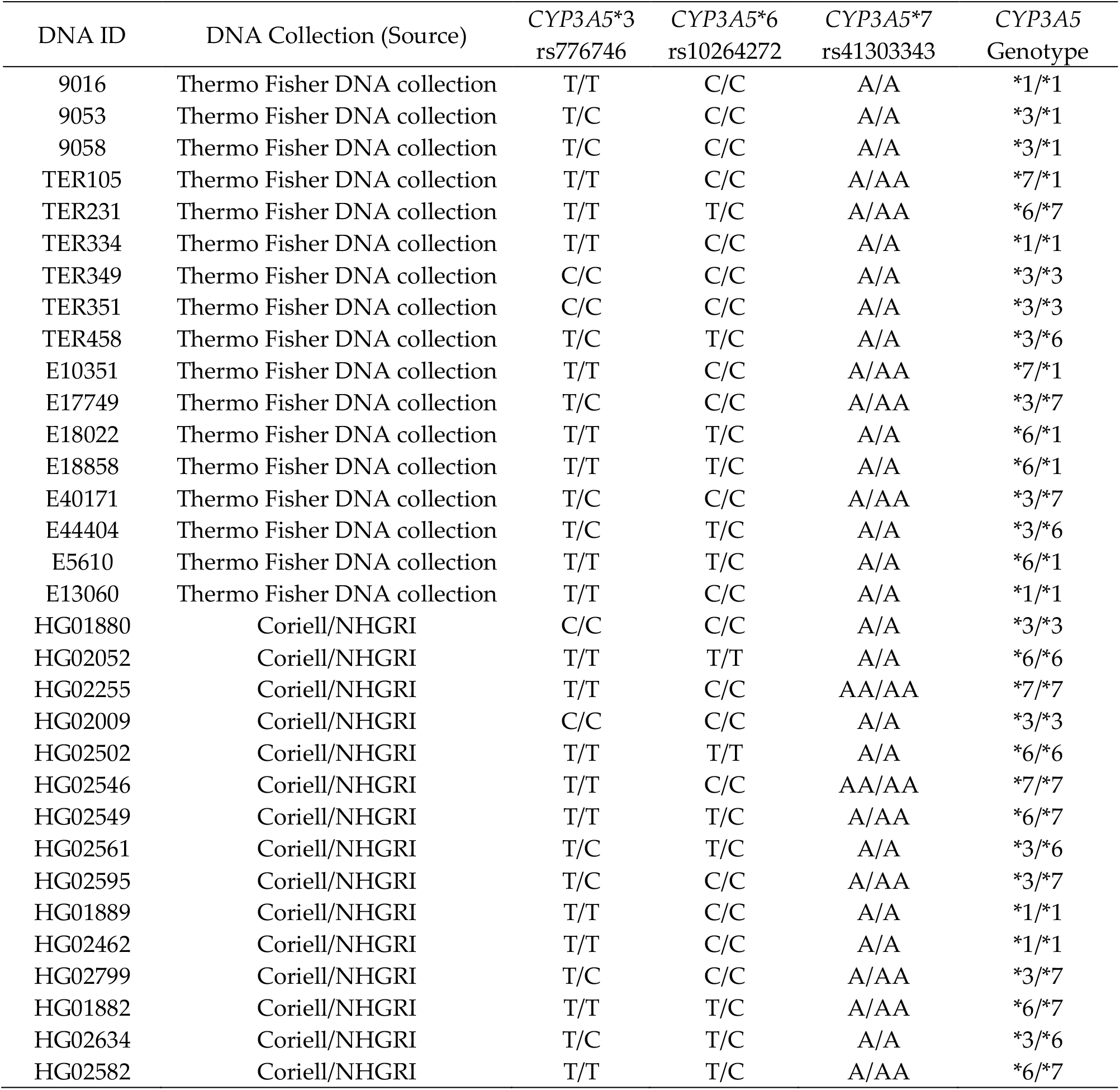
List of reference DNA samples used for verification studies. The allele pair for each SNP is shown.

| DNA ID | DNA Collection (Source) | CYP3A5*3<br>rs776746 | CYP3A5*6<br>rs10264272 | CYP3A5*7<br>rs41303343 | CYP3A5<br>Genotype |
| --- | --- | --- | --- | --- | --- |
| 9016 | Thermo Fisher DNA collection | T/T | C/C | A/A | *1/*1 |
| 9053 | Thermo Fisher DNA collection | T/C | C/C | A/A | *3/*1 |
| 9058 | Thermo Fisher DNA collection | T/C | C/C | A/A | *3/*1 |
| TER105 | Thermo Fisher DNA collection | T/T | C/C | A/AA | *7/*1 |
| TER231 | Thermo Fisher DNA collection | T/T | T/C | A/AA | *6/*7 |
| TER334 | Thermo Fisher DNA collection | T/T | C/C | A/A | *1/*1 |
| TER349 | Thermo Fisher DNA collection | C/C | C/C | A/A | *3/*3 |
| TER351 | Thermo Fisher DNA collection | C/C | C/C | A/A | *3/*3 |
| TER458 | Thermo Fisher DNA collection | T/C | T/C | A/A | *3/*6 |
| E10351 | Thermo Fisher DNA collection | T/T | C/C | A/AA | *7/*1 |
| E17749 | Thermo Fisher DNA collection | T/C | C/C | A/AA | *3/*7 |
| E18022 | Thermo Fisher DNA collection | T/T | T/C | A/A | *6/*1 |
| E18858 | Thermo Fisher DNA collection | T/T | T/C | A/A | *6/*1 |
| E40171 | Thermo Fisher DNA collection | T/C | C/C | A/AA | *3/*7 |
| E44404 | Thermo Fisher DNA collection | T/C | T/C | A/A | *3/*6 |
| E5610 | Thermo Fisher DNA collection | T/T | T/C | A/A | *6/*1 |
| E13060 | Thermo Fisher DNA collection | T/T | C/C | A/A | *1/*1 |
| HG01880 | Coriell/NHGRI | C/C | C/C | A/A | *3/*3 |
| HG02052 | Coriell/NHGRI | T/T | T/T | A/A | *6/*6 |
| HG02255 | Coriell/NHGRI | T/T | C/C | AA/AA | *7/*7 |
| HG02009 | Coriell/NHGRI | C/C | C/C | A/A | *3/*3 |
| HG02502 | Coriell/NHGRI | T/T | T/T | A/A | *6/*6 |
| HG02546 | Coriell/NHGRI | T/T | C/C | AA/AA | *7/*7 |
| HG02549 | Coriell/NHGRI | T/T | T/C | A/AA | *6/*7 |
| HG02561 | Coriell/NHGRI | T/C | T/C | A/A | *3/*6 |
| HG02595 | Coriell/NHGRI | T/C | C/C | A/AA | *3/*7 |
| HG01889 | Coriell/NHGRI | T/T | C/C | A/A | *1/*1 |
| HG02462 | Coriell/NHGRI | T/T | C/C | A/A | *1/*1 |
| HG02799 | Coriell/NHGRI | T/C | C/C | A/AA | *3/*7 |
| HG01882 | Coriell/NHGRI | T/T | T/C | A/AA | *6/*7 |
| HG02634 | Coriell/NHGRI | T/C | T/C | A/A | *3/*6 |
| HG02582 | Coriell/NHGRI | T/T | T/C | A/AA | *6/*7 |

We utilized well-characterized reference samples obtained from the NHGRI Sample Repository for Human Genetic Research (Coriell Institute for Medical Research). *CYP3A5* genotypes for NHGRI samples were identified using 1000 Genomes Project data analyzed using a customized version of the Persephone® genome browser.

Additional DNA samples used in this study were diverse samples from the Terasaki HLA collection maintained in-house. *CYP3A5* genotypes for these samples were obtained using Thermo Fisher SNP Genotyping TaqMan™ Assays.

### Real-Time PCR

We performed real-time PCR reactions using QuantStudio™ 5 and QuantStudio 5 Dx instruments (Thermo Fisher Scientific). Real-time PCR was performed using proprietary primers, probes and thermal cycling conditions (patent application pending).

### Reference assay used for de-novo genotyping of CYP3A5

For primary human samples and other DNAs without known *CYP3A5* genotype, we verified genotype using Taq-Man Assays C 26201809_30 (rs776746), C 30203950_10 (rs10264272) and C 32287188_10 (rs41303343) (Thermo Fisher Scientific), with TaqMan GTXpress™ Master Mix (Thermo Fisher Scientific). Genotypes were analyzed using Design & Analysis Software 2.8.0 (Thermo Fisher Scientific). The manufacturer recommendations for qPCR conditions were used as follows: 95°C for 10 min for enzyme activation followed by 50 cycles of PCR at maximal ramp rate with 95°C for 15 seconds and 63°C for 1 min and 30 seconds.

### Primary human samples

Whole blood and buccal samples were collected by Discovery Life Sciences™ (DLS) from ethnically diverse donors. All samples were deidentified with limited demographic information available. Ethical sample collection and IRB compliance was assured by DLS for all provided materials.

### DNA extraction from buccal swabs

DNA was extracted from buccal swab samples collected with HydraFlock™ 6’’ Sterile Standard Flock Swabs (Puritan™) using the Dynabeads™ SILANE Genomic DNA Extraction kit, QIAamp® DNA Mini kit (Qiagen), and the EZ1&2™ DNA Tissue Kit (Qiagen). The manufacturer’s recommended protocol was followed with the following modifications:

For the Dynabeads™ SILANE Genomic DNA Extraction Kit (referred to hereafter as “Dynabeads”), buccal swabs were broken off into 2 mL LoBind Eppendorf tubes, and 350 µl of TE (10 mM Tris, 1 mM EDTA, pH 8.0) buffer was added to each swab in the Eppendorf tube. Proteinase K (Sigma P2308), RNase A/T1 (Thermo Fisher Scientific EN0551) and Dynabeads lysis buffer treatment took place in the presence of the buccal swab. Following treatment, 500 ul of lysis buffer mixture was transferred to a new tube. 33 µl of Dynabeads MyOne Silane suspension and 265 µl of 100% isopropanol were used.

For the QIAamp DNA Mini kit (Qiagen), 600 µL of PBS was used for initial suspension, 5 uL of RNase A/T1 was added. Final elution was performed in 100 µL of elution buffer.

### DNA extraction from whole blood

In separate studies, blood samples that had been collected in Acid Citrate Dextrose (ACD), ethylenediaminetetraacetic acid (EDTA) and PAXgene® Blood RNA collection tubes were used. Blood was extracted using multiple methods, including Dynabeads, QIAamp DNA Mini kit (Qiagen), QIAamp DNA Blood Mini Kit (Qiagen), EZ1&2 Virus Mini Kit (Qiagen), EZ1&2 DNA Tissue Kit and RSC Whole Blood DNA Kit (Maxwell®). The manufacturer’s recommended protocol was followed with the following modification:

For blood samples extracted with the Dynabeads SILANE Genomic DNA Extraction kit, RNase A/T1 was added to samples following addition of lysis/binding buffer, and final elution was performed in 50 µL.

Whole blood and buccal swab samples were characterized using either a Nanodrop™ spectrophotometer (Thermo Fisher Scientific) or Qubit fluorometer (Thermo Fisher Scientific) using the Qubit dsDNA HS assay kit (Q32854).

### Preparation of Stock Solutions for Interfering Substances Study

For interfering studies performed in blood, Heparin (Tocris Bioscience) was reconstituted in nuclease-free water at a stock concentration of 50 µg/µL. Hemin Chloride (Millipore Sigma) was reconstituted in freshly prepared 0.1 M NaOH at a stock concentration of 1 mg/mL.

### Analytical study design and statistical considerations

Assay characterization tests performed in this study were following CLSI guidelines for genotyping and molecular test methods[63,75].

Concordance was defined as the identity of each allele within the reported allelic pair to the reference typing of the tested sample. The lower bound 95% confidence interval for concordance analysis was calculated by the exact binomial test using R-Studio software

## 4. Patents

Authors are employed by One Lambda Inc., part of Thermo Fisher Scientific Inc., which submitted a patent application covering concepts disclosed in this manuscript relating to TacroType. The submitted application is not published as of the date of this submission.

## Data Availability

Provisional patent application covering concepts disclosed in this manuscript is submitted in the end of 2025

## Author Contributions

“Ab.C” and “An.C” are used to distinguish authors with the same initials. Conceptualization, Ab.C. and An.C.; methodology, Ab.C. and An.C.; software, Ab.C., M.M. and An.C.; validation, Ab.C., Z.W., M.M., JH., LX., S.S. and An.C.; formal analysis, Ab.C., Z.W., M.M., and An.C.; investigation, Ab.C. and An.C.; resources, M.R., A.V., and An.C.; data curation, Ab.C. and An.C.; writing—original draft preparation, Ab.C. and An.C.; writing—review and editing, Ab.C. and An.C. All authors have read and agreed to the published version of the manuscript.

## Institutional Review Board Statement

Not applicable.

## Informed Consent Statement

Additional informed consent is not required for this study as it was obtained by the sample provider, Discovery Life Sciences, who followed IRB protocols with informed consent, and provided de-identified samples.

## Data Availability Statement

The data presented in this study are available on request from the corresponding author upon completion of an ongoing patent application.

## Acknowledgments

The authors thank Gwendolyn George for valuable comments during preparation of the manuscript. The authors thank Brian Yu for technical support in dotting plates for precision studies. The authors thank JJ Chen for support in acquisition of genetic samples from the Coriell Repository. The following cell lines/DNA samples were obtained from the NHGRI Sample Repository for Human Genetic Research at the Coriell Institute for Medical Research: HG01880, HG01882, HG01889, HG02009, HG02052, HG02255, HG02462, HG02502, HG02546, HG02549, HG02561, HG02582, HG02595, HG02634, HG02799.

## Abbreviations

The following abbreviations are used in this manuscript:

CYP3A5: Cytochrome P450 Family 3 Subfamily A Member 5
qPCR: Quantitative Polymerase Chain Reaction
DNA: Deoxyribose Nucleic Acid
Cq: Cycle Quantification
CI: 95% Confidence Interval
FDA: Food and Drug Administration
CPIC: Clinical Pharmacogenetics Implementation Consortium
DPWG: Dutch Pharmacogenetics Working Group
AMP: Association of Molecular Pathology
SNP: Single Nucleotide Polymorphism
QS5: Quantstudio 5
MIQE: Minimum Information for Publication of Quantitative Real-Time PCR Experiments
LDT: Laboratory-Developed Test
EDTA: Ethylenediaminetetraacetic Acid
ACD: Acid Citrate Dextrose
RT: Room Temperature
Pmix: Primer/Probe Mix
NHGRI: National Human Genome Research Institute

